# Eating Disorder Severity and Treatment Outcome Across Race/Ethnicity, Sexual Orientation, and Socioeconomic Status: Intersectional Inequities in a Clinical Sample

**DOI:** 10.1101/2025.05.21.25328100

**Authors:** Catherine R. Drury, Ariel L. Beccia, Samar Shaqour, Megan Riddle, Sasha Gorrell, Simar Singh, Alan Duffy, Philip S. Mehler, Kianna Zucker, Daniel Le Grange, Renee D. Rienecke, Erin E. Reilly

**Affiliations:** Department of Psychiatry and Behavioral Sciences, University of California, San Francisco, 675 18^th^ Street, San Francisco, CA 94143, USA; Division of Adolescent and Young Adult Medicine, Boston Children’s Hospital, Boston, MA, USA; Department of Pediatrics, Harvard Medical School, Boston, MA, USA; Department of Epidemiology, Harvard T.H. Chan School of Public Health, Boston, MA, USA; Eating Recovery Center and Pathlight Mood & Anxiety Center, Denver, CO, USA; ACUTE Center for Eating Disorders at Denver Health, Denver, CO, USA; University of Colorado School of Medicine, Aurora, CO, USA; Department of Psychiatry and Behavioral Sciences, The University of Chicago, IL, USA (Emeritus); Department of Psychiatry and Behavioral Sciences, Northwestern University, Chicago, IL, USA

## Abstract

**Background:** Marginalized populations experience increased eating disorder (ED) risk and encounter significant barriers to treatment. Intersectionality provides a framework for understanding how systemic oppression contributes to inequities in EDs; however, intersectional approaches have yet to be applied to a clinical ED sample. The current study examined inequities in ED severity and treatment outcome across the intersections of race/ethnicity, sexual orientation, and socioeconomic status (SES).

**Methods:** Adult women (*N=*3,016; *M*=27.2 years) with transdiagnostic EDs presenting to affiliated treatment sites across the United States completed the Eating Disorder Examination-Questionnaire (EDE-Q) at admission and discharge. Race/ethnicity and sexual orientation were self-reported; SES was measured using the area deprivation index of participants’ neighborhoods. Multilevel Analysis of Individual Heterogeneity and Discriminatory Accuracy (MAIHDA) was used to estimate baseline EDE-Q global score; change in EDE-Q global score and binge eating, self-induced vomiting, laxative use, and driven exercise frequency from admission to discharge; and reason for discharge (routine or non-routine) across intersectional subgroups.

**Results:** In this sample of women with access to treatment, MAIHDA models predicted higher baseline levels of overall ED pathology among sexual minorities (predicted *M*=4.10). There were few differences in ED symptom improvement across intersectional subgroups, with some small yet potentially meaningful inequities. Racially/ethnically minoritized subgroups were slightly less likely to complete treatment (predicted percent non-routine discharge=41.50%).

**Conclusions:** Future research should build on these findings by analyzing other dimensions of inequity (e.g., gender, weight status, disability status) to further characterize and address intersecting systems of oppression that disparately influence ED outcomes.

## Introduction

Eating disorders (EDs) cause significant suffering and impairment via their impact on physical and psychological health and quality of life (van Hoeken & Hoek, 2020). Although EDs occur across age, race, ethnicity, gender, sexual orientation, and socioeconomic status (SES; Diemer et al., 2018; Grammer et al., 2021; Huryk et al., 2021; Rodgers et al., 2018), ED research has historically overrepresented a particular patient demographic (i.e., White, middle- to upper-class, cisgender women), due in part to longstanding, erroneous stereotypes about who is affected (Halbeisen et al., 2022). This imbalance has created a circular, self-perpetuating problem, whereby individuals who fit this demographic are more likely to be identified as having an ED (Gordon et al., 2006; Sala et al., 2024) and therefore more likely to receive ED treatment and participate in research (Sonneville & Lipson, 2018; Penwell et al., 2024). Thus, while the field continues to uncover heterogeneous presentations and develop tailored treatments (Anderson et al., 2023), ED pathology and treatment response among historically marginalized populations remain poorly characterized (Burnette et al., 2022). To encourage the development of inclusive assessment and treatment approaches, additional research on how EDs present and progress among marginalized populations is urgently needed (Goel et al., 2022; Halbeisen et al., 2022).

In response to calls to dismantle ED stereotypes (Halbeisen et al., 2022), an increasing number of community-based studies have examined the distribution of ED pathology within and between social groups. Critically, this emerging research has found that marginalized populations may be disproportionately affected by EDs. For example, forms of adversity/disadvantage such as discrimination and food insecurity increase ED risk (Hazzard et al., 2020; Mason et al., 2021). Additionally, sexual and gender minorities exhibit higher rates of EDs and ED pathology than heterosexual and cisgender people (Nagata et al., 2024), and several studies suggest a heightened prevalence of EDs and ED behaviors among socially minoritized racial, ethnic, and multiracial groups (Burke et al., 2021; Gordon et al., 2024; Simone et al., 2022).

Intersectionality provides a more nuanced framework for examining health inequities, including in ED presentation and treatment outcomes. Based in Black feminist scholarship (Crenshaw, 1993), intersectionality recognizes that systems of power and oppression embedded in society (e.g., structural racism, structural sexism) intersect, or mutually constitute one another, differentially shaping lived experiences according to one’s unique constellation of social identities (e.g., race/ethnicity, gender). Thus, intersectionality recognizes the variability in risk and resilience within heterogeneous populations such as “people of color” or “women” (Bowleg, 2012). Regarding health inequities, it is increasingly understood that unjust societal systems and structures directly influence disease risk and access to care (Bailey et al., 2017; Homan, 2019). Examining health inequities along a single axis (e.g., by gender) or only among multiply marginalized populations (i.e., not including people experiencing multiple forms of privilege or both privilege and disadvantage) is therefore insufficient (Cole, 2009; Evans et al., 2018). Instead, an intersectional perspective requires consideration of the ways by which multiple inequities (as proxies for experience with systems of power and oppression) interact to shape risk for a given outcome across intersectional social groups.

Intersectional research approaches hold promise for illuminating and addressing persistent inequities in EDs (Burke et al., 2020). To date, only a handful of studies (Beccia et al., 2019, 2021; Billman Miller et al., 2024; Burke et al., 2023; Egbert et al., 2024; Gordon et al., 2024) have considered ED pathology from an intersectional framework – even fewer have examined the interaction of more than two axes of inequity. Consistent with intersectionality, this burgeoning research area has found particularly high ED prevalence among multiply marginalized populations. For example, Burke and colleagues (2023) studied EDs at the intersection of SES, gender identity, sexual orientation, and race/ethnicity, and found disproportionately high prevalence among Hispanic/Latine, sexual minority men, and lower-SES women, with considerable heterogeneity across all groups. Likewise, Beccia and colleagues (2021) examined ED pathology and diagnosis across the intersections of gender identity and expression, sexual orientation, and weight status and identified several multiply marginalized populations with heightened ED prevalence, including gender non-conforming, sexual minority, and larger-bodied girls and women. Notably, these inequities occurred over and above individual-level differences, underscoring the importance of adopting an intersectional perspective that explicitly considers structural influences.

All existing intersectional ED research has been conducted in non-clinical, community samples, leaving unstudied the ways in which structural and systemic factors that contribute to inequities in ED onset may also influence variability in illness course. For example, given that marginalized individuals are more likely to encounter barriers to specialized ED care (Lane et al., 2020; Moreno et al., 2023; Penwell et al., 2024), including delays in ED identification (Gordon et al., 2006; Sala et al., 2024; Sonneville & Lipson, 2018), there may be disparities in ED severity at initial presentation to treatment. Moreover, considering certain social groups have been underrepresented in the development of ED treatments and treatment programs (Burnette et al., 2022; Halbeisen et al., 2022), there may be inequities in treatment response across intersectional social group as well.

The current study sought to fill these gaps by investigating intersectional inequities within a transdiagnostic sample of adults treated for EDs at intensive ED treatment centers. Specifically, we examined the intersection of race/ethnicity, sexual orientation, and SES relative to (a) baseline level of ED pathology and (b) treatment outcome, as measured by change in ED pathology from admission to discharge and reason for discharge from treatment. Based on prior work, we hypothesized that multiply marginalized patients would exhibit higher levels of ED pathology at baseline and less improvement in symptoms than multiply privileged patients, and be less likely to complete a full course of treatment.

## Method

### Participants and Procedure

Participants were adults with transdiagnostic EDs admitted to a multisite ED treatment facility in the United States (US) October 2020-September 2023. Treatment sites spanned 24 locations and included inpatient units, non-hospital-based residential treatment centers, partial hospitalization programs (PHP), and intensive outpatient programs (IOP). All sites utilized a singular treatment approach, detailed in the Supplementary Material.

Sociodemographic information and ED diagnoses were determined during semi-structured interviews administered by licensed clinicians and informed by *DSM-5* (American Psychiatric Association, 2013) criteria. Upon admission, patients were approached regarding study participation and provided informed consent before completing self-report questionnaires, which were readministered at treatment discharge. Study procedures were approved by the Salus Institutional Review Board.

Of the 3,798 patients who provided informed consent and completed outcome measures at baseline, 82.2% were cisgender women. Patients who identified as cisgender men (7.0%), transgender (1.8%), non-binary (6.0%), or did not disclose a gender identity (3.1%) were excluded from the current study due to having insufficient sample sizes for intersectional analyses. Additional exclusion criteria included missing both race/ethnicity and sexual orientation data (2.8%), resulting in a final sample size of 3,016 cisgender women with EDs.

### Measures

#### Social Identities

Consistent with intersectionality, we emphasize that the social identities and labels used in this study stand as proxies for experiences with systems of oppression and other social processes (e.g., structural racism, classism) that may differentially influence ED severity and treatment response. For each of the social identities examined, decisions were made to categorize participants to allow sufficient sample sizes for intersectional analysis, while preserving nuance and variability within the data as much as possible. Although similar classifications have been used in prior intersectionality-based studies (e.g., Beccia et al., 2019; Forrest et al., 2023), this is a notable limitation addressed further in the Discussion.

##### Race/Ethnicity

During initial clinical interview, clinicians asked participants an open question regarding their race/ethnicity, selected one of seven options that combined race/ethnicity into a single variable, and confirmed their selection with participants: American Indian/Native American or Alaska Native; Asian; Black/African American; Hispanic/Latine; Native Hawaiian/Pacific Islander; White; or Other, which included bi/multiracial patients. Some participants reported a more specific race/ethnicity (e.g., Indian), which was noted in a comments section. Specific geographic nationalities (e.g. Japanese) were then converted to the relevant classification (e.g. Asian); where such a conclusion could not be made, Other was used to avoid assumptions about racial/ethnic identity. Due to sample size limitations, American Indian/Native American or Alaska Native and Native Hawaiian/Pacific Islander were collapsed into a single category, termed Indigenous.

##### Sexual Orientation

Sexual orientation was also assessed during initial clinical interview as part of a dual, open-ended question regarding both sexual orientation and relationship status. Because clinicians often recorded patients’ relationship status without noting their sexual orientation, there was a high degree of missingness in sexual orientation data (54.5%). Responses comprised 11 categories: asexual, bisexual, gay, lesbian, mostly heterosexual, pansexual, queer, questioning, heterosexual, multiple identities, and other. Based on documented differences in mental health outcomes between gay or lesbian and bisexual people (Chan et al., 2020; Pitman et al., 2022), the following collapsed categories were used: heterosexual; gay or lesbian; bisexual, pansexual, or queer; and “other,” which included patients who identified as asexual, mostly heterosexual, multiple identity, other, and questioning.

##### Socioeconomic Status

We operationalized SES as the area deprivation index (ADI; Kind & Buckingham, 2018) of the home neighborhoods in which participants resided at the time of admission. Participants’ zip codes were used to calculate ADI based on 5-year (2018-2022) American Community Survey data regarding the income, education, employment, and housing quality of each area. We mapped zip codes to zip code tabulation areas (ZCTA) using a crosswalk table (Human Resources and Service Administration, 2020); the ‘sociome’ package (Krieger et al., 2023) for R was used to calculate ADI. Because our sample represented all 50 states plus the District of Columbia, the reference area was set to all ZCTA in the US such that ADI scores were assigned relative to all other US ZCTA. By definition, the ADI has an *M* of 100 and an *SD* of 20, with higher ADI scores representing greater socioeconomic disadvantage. For the current study, ADI scores were converted to a binary variable (0=higher SES, 1=lower SES) divided along the median score (ADI=82.75), as in prior work (e.g., Deng et al., 2022; Hufnagel et al., 2021).

#### Eating Disorder Pathology

The Eating Disorder Examination-Questionnaire (EDE-Q; Fairburn & Beglin, 1994, 2008) is a 28-item self-report measure of ED pathology over the past 28 days. The measure generates four subscales, the average of which yields a global score. Items not included in the global score assess the frequency of binge eating episodes, self-induced vomiting, laxative use, and driven exercise. In the current study, internal consistency for the EDE-Q global score was Cronbach’s α=.91 at admission and α=.92 at discharge.

#### Discharge Status

Treatment center staff were trained to identify and record which of the following categories best captured patients’ reason for discharge: routine (i.e., deemed appropriate by patient’s care team based on symptom improvement or need for a different care setting), administrative (i.e., requested by care team due to unacceptable behavior or poor program fit), resource constraint (i.e., insurance denying/ending coverage, patient unable to pay for treatment), patient request (i.e., without medical risk but prior to the care team recommending discharge), against medical advice (i.e., patient leaving treatment when not advised due to medical risk), and COVID-19 (i.e., due to contracting COVID or fear of contracting COVID). Similar to prior work (Gorrell et al., 2022), discharge status was operationalized as a binary construct (0=routine, 1=non-routine) to examine intersectional inequities in likelihood of completing a full course of treatment.

### Statistical Analyses

Multilevel Analysis of Individual Heterogeneity and Discriminatory Accuracy (MAIHDA) was used to examine inequities in ED outcomes at the intersection of race/ethnicity, sexual orientation, and SES. Considered best practice in quantitative intersectionality research (Evans et al., 2018; Merlo, 2018), MAIHDA is a theoretically driven, descriptive approach that involves fitting multilevel models in which individuals at level 1 are nested within intersecting subgroups or “social strata” at level 2. Stratum inequities are therefore examined not only in terms of average differences between stratum, but also as they relate to the total outcome heterogeneity at the stratum (versus individual) level, quantifying a “contextual effect” (Merlo, 2018). By avoiding a purely additive approach (e.g., single-level regression with interaction terms), MAIHDA further aligns with intersectionality in that each social stratum is uniquely considered to hold advantages and disadvantages relative to the outcome. A published tutorial for MAIHDA (Evans et al., 2024) informed our analyses and interpretation.

We first examined sample sizes across social strata (Table S1). Although MAIHDA accommodates small strata sample sizes by providing more conservative estimates via shrinkage, current guidelines recommend ≥20 participants in at least 80% of all strata (Evans et al., 2018, 2024). Thus, due to the large amount of missing sexual orientation data constraining strata sample sizes, we chose to conduct two separate analyses across the intersection of: 1) race/ethnicity and SES, and 2) sexual orientation and SES. Outcomes were EDE-Q global score at admission; change in EDE-Q global score, change in binge eating episode frequency, change in self-induced vomiting frequency, change in laxative use frequency, and change in driven exercise episode frequency from admission to discharge; and reason for discharge.

For each of these 14 outcomes, we fit a two-level multilevel *simple intersectional model* with random intercepts for social strata. From each model, we calculated the *variance partition coefficient (VPC)* as a measure of the proportion of total sample variance in the outcome that can be attributed to between-stratum variance, and predicted outcomes for each social stratum. As a descriptive analysis, MAIHDA does not focus on significance testing; instead, results were interpreted in the context of the VPCs and between-stratum differences in predicted outcomes. Although we planned to fit intersectional interaction models that adjusted for the additive effects of social identity (Evans et al., 2024), low levels of heterogeneity in the simple intersectional models precluded such analyses.

Data were analyzed in R version 4.4.1 (2023). The ‘cmdstanr’ package (Gabry et al., 2024) was used to fit the MAIHDA models using Bayesian Markov chain Monte Carlo estimation methods, with vague priors, a burn-in period of 1,000 iterations, and a total of 8,000 iterations for each model (Evans et al., 2024). Posterior samples were checked using R-hat and effective sample size, and visual assessments of trace plots and autocorrelation plots. Consistent with descriptive epidemiologic approaches, we did not include any adjustment variables in our models that might obscure the magnitude of inequities across social strata (Fox et al., 2022; Lesko et al., 2022). For example, prior intersectional studies of ED risk have adjusted for age to account for variability in ED risk across the age spectrum; however, given that the current study used a clinical sample, differences in outcomes across age groups may reflect barriers to care that lead to marginalized groups accessing treatment at a later age. Thus, adjusting for age would adjust away the inequitable systems that influence treatment access and clinical outcomes. Although we explored adjusting for baseline scores in the change score analyses, this can also introduce bias when the exposure (e.g., oppression) also predicts baseline level of the outcome (Glymour et al., 2005). As the pattern of findings was similar, with comparable VPC values for all models and reduced heterogeneity across strata (Table S2), unadjusted results are presented.

## Results

Sociodemographic and clinical characteristics for the entire sample (*N*=3,016) are shown in Table 1. Most participants were White (84.2%) and heterosexual (66.8%), with an average ADI score less than the average for all US ZCTAs (i.e., less socioeconomic disadvantage). Most participants had a diagnosis of anorexia nervosa (45.9%) or other specified feeding or eating disorder (33.7%).

**Table 1.** Participant Demographic and Clinical Characteristics.

|  | *n* | % |  |
| --- | --- | --- | --- |
| Level of care at admission |  |  |  |
| Inpatient | 610 | 20.2 |  |
| Residential | 1,007 | 33.4 |  |
| PHP | 757 | 25.1 |  |
| IOP | 168 | 5.6 |  |
| Virtual IOP | 473 | 15.7 |  |
| ED diagnosis |  |  |  |
| AN-R | 908 | 30.1 |  |
| AN-BP | 478 | 15.8 |  |
| BN | 234 | 7.8 |  |
| BED | 196 | 6.5 |  |
| ARFID | 176 | 5.8 |  |
| OSFED | 1,017 | 33.7 |  |
| UFED | 7 | 0.2 |  |
| Race/Ethnicity |  |  |  |
| Indigenous<sup>a</sup> | 13 | 0.4 |  |
| Asian | 95 | 3.2 |  |
| Black/African American | 96 | 3.2 |  |
| Hispanic/Latine | 163 | 5.5 |  |
| White | 2,517 | 84.2 |  |
| Bi/multiracial | 105 | 3.5 |  |
| Sexual Orientation |  |  |  |
| Heterosexual | 917 | 66.8 |  |
| Gay or lesbian | 86 | 6.3 |  |
| Bisexual, pansexual, or<br>queer | 282 | 20.5 |  |
| Other<sup>b</sup> | 88 | 6.4 |  |
| Reason for discharge |  |  |  |
| Routine | 1,844 | 61.1 |  |
| Non-routine | 1,172 | 38.9 |  |
|  | *M* | *SD* | Range |
| Age (years) | 27.21 | 10.49 | 15.00-68.00 |
| Area deprivation index (ADI)<sup>c</sup> | 82.72 | 18.66 | 21.17-140.58 |
| Total length of stay (days) | 66.73 | 48.73 | 1.00-738.00 |
| %EBW at admission | 103.64 | 38.82 | 38.13-425.47 |
| ED symptoms at admission |  |  |  |
| EDE-Q global score | 3.73 | 1.57 | 0.00-6.00 |
| Binge eating episodes | 5.50 | 15.08 | 0.00-500.00 |
| Self-induced vomiting | 6.37 | 21.56 | 0.00-500.00 |
| Laxative use | 2.10 | 11.10 | 0.00-500.00 |
| Compulsive exercise | 5.69 | 10.82 | 0.00-300.00 |

*Note.* $N = 3,016$ . Percents are valid percents. PHP = partial hospitalization program; IOP = intensive outpatient program; ED = eating disorder; AN-R = anorexia nervosa, restricting type; AN-BP = anorexia nervosa, binge-purge type; BN = bulimia nervosa; BED = binge eating disorder; ARFID = avoidant/restrictive food intake disorder; OSFED = other specified feeding or eating disorder; UFED = unspecified feeding or eating disorder; EBW = expected body weight; EDE-Q = Eating Disorder Examination-Questionnaire; SES = socioeconomic status.
<sup>a</sup> Indigenous includes American Indian/Native American or Alaska Native and Native Hawaiian/Pacific Islander participants.
<sup>b</sup> Other sexual orientation includes participants who identified as asexual, mostly straight, multiple identity, other, and questioning.
<sup>c</sup> ADI scores were assigned relative to all US zip code tabulation areas ( $M = 100$ , $SD = 20$ ). Higher ADI scores represent greater socioeconomic disadvantage.

Similar to other studies of ED higher levels of care (e.g., Brown et al., 2018; Reilly et al., 2020), a large number of participants (46.0%) were missing EDE-Q data at discharge and therefore excluded from change score analyses. Participants with missing data were older, *p=*.034, had a shorter length of stay, *p*<.001, lower EDE-Q global scores, *p=*.014, and were at a higher percent of expected body weight at admission, *p*<.001. Indigenous, Hispanic/Latine, and bi/multiracial participants had a larger proportion of missingness than White participants, *p*=.009. There was also a larger proportion of missingness among participants admitted to PHP or IOP, with a non-routine discharge, and with an ED diagnosis other than anorexia nervosa (all *p*s<.001). Participants with and without missing EDE-Q data did not significantly differ in sexual orientation or ADI.

Table 2 provides a summary of strata sample sizes, which were sufficient for both models, with ≥20 participants in 83% and 100% of strata. When the sample was reduced to include only participants with EDE-Q data at discharge for the change score analyses, there were ≥20 participants in 75% of strata for the race/ethnicity and SES model and 100% of strata for the sexual orientation and SES model.

**Table 2.** Sample Sizes of Intersectional Social Strata.

| Strata defined by race/ethnicity and SES ($n = 12$) | | |
| --- | --- | --- |
| Sample size per stratum | Number of strata | % of strata |
| 100 or more | 2 | 16.7 |
| 50 or more | 7 | 58.3 |
| 30 or more | 10 | 83.3 |
| 20 or more | 10 | 83.3 |
| 10 or more | 11 | 91.7 |
| Less than 10 | 1 | 8.3 |
| Strata defined by sexual orientation and SES ($n = 8$) | | |
| Sample size per stratum | Number of strata | % of strata |
| 100 or more | 4 | 50.0 |
| 50 or more | 4 | 50.0 |
| 30 or more | 8 | 100.0 |
| 20 or more | 8 | 100.0 |
| 10 or more | 8 | 100.0 |
| Less than 10 | 0 | 0.0 |

*Note.* SES = socioeconomic status as measured by area deprivation index (ADI) divided along the median (ADI=82.75).

Results for all simple intersectional MAIHDA models are shown in Table 3 (baseline-adjusted model results in Table S2); Figures 3 and 4 show the stratum-specific predicted values for each outcome. Full summaries of strata sample sizes, observed and predicted values, and baseline-adjusted predicted values are provided in the Supplementary Material (Tables S3-S9). For ED severity at admission, VPCs indicated that only 0.2% of the total variation in ED pathology was attributable to stratum-level differences across race/ethnicity and SES, while 2.1% was attributable to stratum-level differences across sexual orientation and SES, over and above differences between individuals. Table A3 shows the stratum-specific predicted EDE-Q global scores at admission, which were similar across social strata in the race/ethnicity and SES model (range: 3.66-3.74). Strata including sexual minorities had slightly higher predicted levels of ED pathology at admission than those including heterosexual patients (range: 3.71-4.10), with lower-SES gay or lesbian patients having the highest predicted values.

**Table 3.**
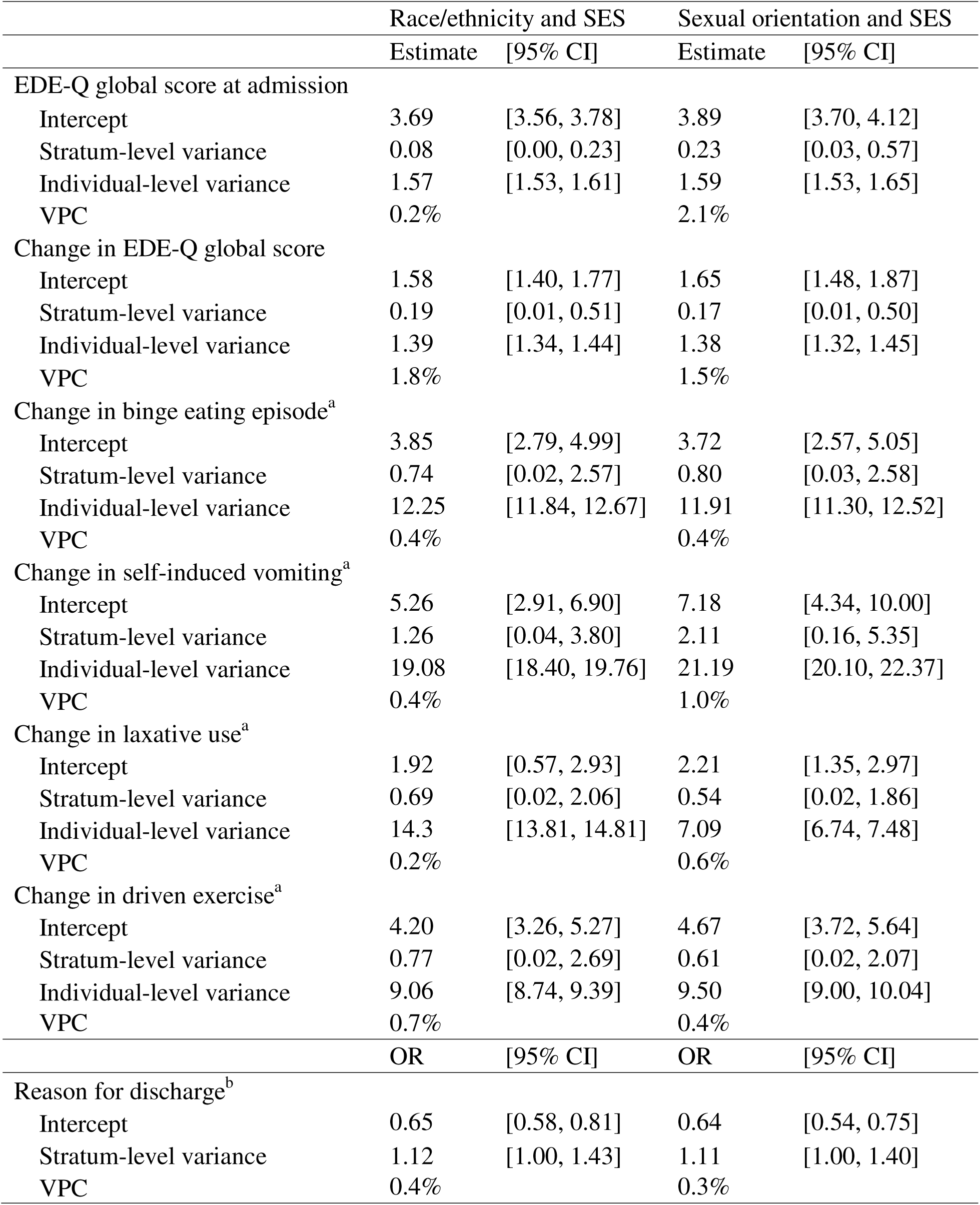
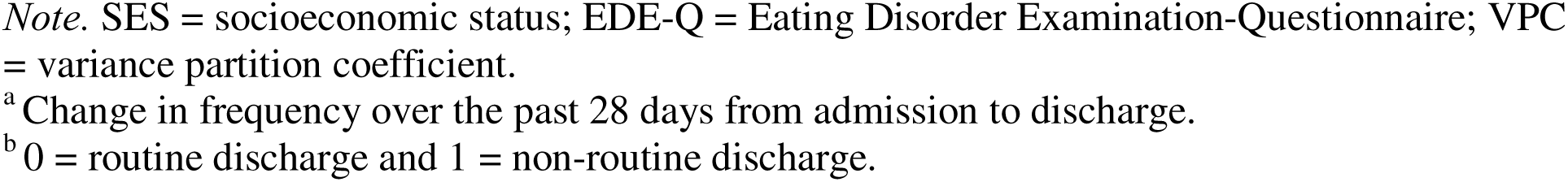
Parameter Estimates for Simple Intersectional Models.

|  | Race/ethnicity and SES |  | Sexual orientation and SES |  |
| --- | --- | --- | --- | --- |
|  | Estimate | [95% CI] | Estimate | [95% CI] |
| EDE-Q global score at admission |  |  |  |  |
| Intercept | 3.69 | [3.56, 3.78] | 3.89 | [3.70, 4.12] |
| Stratum-level variance | 0.08 | [0.00, 0.23] | 0.23 | [0.03, 0.57] |
| Individual-level variance | 1.57 | [1.53, 1.61] | 1.59 | [1.53, 1.65] |
| VPC | 0.2% |  | 2.1% |  |
| Change in EDE-Q global score |  |  |  |  |
| Intercept | 1.58 | [1.40, 1.77] | 1.65 | [1.48, 1.87] |
| Stratum-level variance | 0.19 | [0.01, 0.51] | 0.17 | [0.01, 0.50] |
| Individual-level variance | 1.39 | [1.34, 1.44] | 1.38 | [1.32, 1.45] |
| VPC | 1.8% |  | 1.5% |  |
| Change in binge eating episode<sup>a</sup> |  |  |  |  |
| Intercept | 3.85 | [2.79, 4.99] | 3.72 | [2.57, 5.05] |
| Stratum-level variance | 0.74 | [0.02, 2.57] | 0.80 | [0.03, 2.58] |
| Individual-level variance | 12.25 | [11.84, 12.67] | 11.91 | [11.30, 12.52] |
| VPC | 0.4% |  | 0.4% |  |
| Change in self-induced vomiting<sup>a</sup> |  |  |  |  |
| Intercept | 5.26 | [2.91, 6.90] | 7.18 | [4.34, 10.00] |
| Stratum-level variance | 1.26 | [0.04, 3.80] | 2.11 | [0.16, 5.35] |
| Individual-level variance | 19.08 | [18.40, 19.76] | 21.19 | [20.10, 22.37] |
| VPC | 0.4% |  | 1.0% |  |
| Change in laxative use<sup>a</sup> |  |  |  |  |
| Intercept | 1.92 | [0.57, 2.93] | 2.21 | [1.35, 2.97] |
| Stratum-level variance | 0.69 | [0.02, 2.06] | 0.54 | [0.02, 1.86] |
| Individual-level variance | 14.3 | [13.81, 14.81] | 7.09 | [6.74, 7.48] |
| VPC | 0.2% |  | 0.6% |  |
| Change in driven exercise<sup>a</sup> |  |  |  |  |
| Intercept | 4.20 | [3.26, 5.27] | 4.67 | [3.72, 5.64] |
| Stratum-level variance | 0.77 | [0.02, 2.69] | 0.61 | [0.02, 2.07] |
| Individual-level variance | 9.06 | [8.74, 9.39] | 9.50 | [9.00, 10.04] |
| VPC | 0.7% |  | 0.4% |  |
|  | OR | [95% CI] | OR | [95% CI] |
| Reason for discharge<sup>b</sup> |  |  |  |  |
| Intercept | 0.65 | [0.58, 0.81] | 0.64 | [0.54, 0.75] |
| Stratum-level variance | 1.12 | [1.00, 1.43] | 1.11 | [1.00, 1.40] |
| VPC | 0.4% |  | 0.3% |  |

*Note.* SES = socioeconomic status; EDE-Q = Eating Disorder Examination-Questionnaire; VPC = variance partition coefficient.
<sup>a</sup> Change in frequency over the past 28 days from admission to discharge.
<sup>b</sup> 0 = routine discharge and 1 = non-routine discharge.

Regarding change in ED symptoms from admission to discharge, heterogeneity across social strata for all outcomes was low, with small inequities in symptom reductions. All strata seemed to similarly benefit from treatment in terms of overall ED pathology (predicted change score range: 1.42-1.83; VPCs=1.5-1.8%). Across both the race/ethnicity and SES model and the sexual orientation and SES model, strata with lower SES tended to experience greater reductions in ED pathology than strata with higher SES, with lower-SES Hispanic/Latine and gay or lesbian patients demonstrating the greatest improvements (Table S4). Change in overall ED pathology was lowest among strata including higher-SES Asian and heterosexual patients.

While reductions in binge eating episode frequency (Table S5) were similar across social strata (predicted change score range: 3.29-4.53; VPCs=0.4%), small inequities were found for self-induced vomiting frequency (predicted change score range: 4.79-8.85; VPCs=0.4-1.0%), with a complex patterning across both models (Table S6). Although several strata including lower-SES patients appeared to experience smaller improvements in self-induced vomiting than their higher-SES counterparts, lower-SES Black/African American and bisexual, pansexual, or queer patients demonstrated the greatest reductions in self-induced vomiting frequency in each model. Lower-SES bi/multiracial patients had the smallest predicted change scores in the race/ethnicity and SES model, while heterosexual patients showed the least improvement in self-induced vomiting relative to sexual minorities.

Similarly, low levels of heterogeneity were found for change in laxative use (predicted change score range: 1.80-2.39; VPCs=0.2-0.6%) and driven exercise frequency (predicted change score range: 3.58-5.02; VPCs=0.4-0.7%). Social strata with White patients experienced greater reductions in laxative use than those with racially/ethnically minoritized patients (Table S7). Lower-SES patients whose sexual orientation was categorized as “other” demonstrated the least improvement in laxative use and driven exercise frequency (Table S8).

Results from the reason for discharge models are shown in Tables 3 and S9. A small proportion of the variation in reason for discharge was attributable to stratum-level differences (predicted percent non-routine discharge range: 38.12-41.50%; VPCs=0.3-0.4%) and suggested a greater likelihood of a non-routine discharge among strata that included racially/ethnically minoritized patients, with Indigenous patients having the highest predicted prevalence. Lower-SES White and heterosexual patients were least likely to have a non-routine discharge.

## Discussion

The current study is the first to apply an intersectional approach to a clinical ED sample by examining and quantifying inequities in ED severity and treatment outcome across the intersections of race/ethnicity, sexual orientation, and SES. At admission to specialized ED treatment, sexual minorities reported higher levels of ED pathology than heterosexual patients, with lower-SES gay or lesbian patients endorsing the most severe symptoms. This finding is consistent with the disproportionate rates of ED pathology in community-based sexual minority samples (Calzo et al., 2017; Kamody et al., 2019) as a result of the discrimination and stigma experienced by these individuals (Meyer, 2003). The similar levels of pathology found across racial/ethnic and SES groups further discredits the myth that EDs predominantly affect affluent, White populations. A growing body of work has found increased ED risk among racially/ethnically minoritized and lower-SES people (e.g., Burke et al., 2021; Egbert et al., 2024; Gordon et al., 2024). The current study builds on this literature to suggest that diverse social groups who experience EDs and present to treatment exhibit comparable levels of illness severity, further increasing their risk for the physiological and psychological consequences of EDs.

Across ED symptoms, there was minimal heterogeneity in change from admission to discharge, suggesting that overall, patients benefited similarly from specialized treatment, consistent with prior studies of ED treatment outcomes across sexual and racial/ethnic identities (e.g., Donahue et al., 2020; Lydecker et al., 2019). Within the small treatment inequities observed in the current study, the patterning across intersectional social groups was complex and seemingly inconsistent; however, several findings may highlight important directions for future investigation. First, lower-SES patients experienced more improvement in overall ED pathology than higher-SES patients. However, some strata including lower-SES patients (e.g., White, bi/multiracial, “other” sexual orientation) experienced inequities in reduction of behavioral symptoms, particularly self-induced vomiting. Prior work has found that lower-SES people who experience food insecurity report more frequent binge eating and purging symptoms than those with reliable food access (Abene et al., 2023; Hazzard et al., 2020). In higher level of care treatment for EDs, food is provided for patients to varying degrees; however, this may magnify fluctuations in food availability (i.e., “feast-or-famine” cycles; Hazzard et al., 2020) hypothesized to exacerbate binge-purge symptoms. Thus, while our results align with qualitative accounts that experiences of oppression and deprivation impact ED treatment (Calzo et al., 2025; Frayn et al., 2022), more research is needed to better understand the treatment experiences of lower-SES patients, including in higher levels of care. Finally, the fact that some minoritized groups with lower SES (e.g., Black/African American; bisexual, pansexual, or queer) experienced the greatest improvement in this area emphasizes the advantages of examining multiple social identities simultaneously, as opposed to a unidimensional approach (e.g., comparing lower-SES to higher-SES), which would have obscured this complex patterning of outcomes.

Although variability in discharge reason across intersecting social groups was also low, small inequities were found among strata including racially/ethnically minoritized patients, who were generally less likely to complete treatment than White patients. A non-routine discharge likely involved leaving treatment prior to symptom remission, potentially impeding full recovery and thus contributing to further inequities in outcomes. Critically, research on ED treatment among racially/ethnically minoritized populations remains limited. While a handful of studies have found that neither race nor ethnicity are significant predictors of treatment effectiveness in ED intervention trials (Thompson-Brenner et al., 2013; Linardon et al., 2017), racially/ethnically minoritized groups were more likely to drop out of treatment (Thompson-Brenner et al., 2013). Moreover, a growing body of work, including qualitative research, has highlighted the importance of incorporating culture and experiences of racism and acculturative stress into ED conceptualizations and interventions (Acle et al., 2021). To address inequities in treatment retention, clinicians and ED treatment programs must also consider how they may inadvertently perpetuate systems of power and oppression through implicit biases and the content and structure of services. Such work is urgent if we are to provide accessible, inclusive treatment that makes recovery possible for patients of all racial, ethnic, and cultural identities.

The primary strength of this study is its application of an intersectional approach to describing a large, clinical sample of individuals with transdiagnostic EDs treated under the same treatment paradigm. While addressing a critical gap in the ED literature, this study has several limitations that should be considered for further study. First, although participants represented a diverse range of racial/ethnic, sexual, and socioeconomic identities, the majority were White, heterosexual, cisgender women, and lived in less socioeconomically disadvantaged areas. As EDs occur across races/ethnicities, sexual orientations, genders, and socioeconomic backgrounds and disproportionately impact marginalized groups (Burke et al., 2023; Diemer et al., 2018; Grammer et al., 2021; Rodgers et al., 2018), the limited representation of minoritized and marginalized patients may be attributed to this being a sample with access to ED treatment primarily covered by private health insurance. Due to structural racism and other systems of power and oppression, individuals whose identities do not align with historical ED stereotypes face substantial barriers (e.g., financial, stigma, delayed diagnosis) to accessing specialized care (Penwell et al., 2024). Regarding the impact on our findings, social strata including marginalized patients had smaller sample sizes and, therefore, potentially less accurate predicted values that may have underestimated inequities (Evans et al., 2018, 2024). Future research can recruit clinical samples more representative of the diverse population affected by EDs (e.g., Beccia et al., 2021; Burke et al., 2023), including patients with public health insurance and those without insurance (Accurso et al., 2021).

Another significant limitation is the amount of missingness in the data which, though similar to other research in ED higher levels of care (e.g., Reilly et al., 2020), may have impacted results in several ways, including further constraining the sample sizes of strata that included racially/ethnically minoritized patients in change score analyses and limiting generalizability primarily to more severe ED presentations. The large amount of missing sexual orientation data precluded examining intersections of race/ethnicity, sexual orientation, and SES simultaneously, which would provide a clearer picture of the patterning of inequities across all intersecting subgroups. Other assessment-based limitations include collapsing racial/ethnic and sexual orientation categories, which does not reflect the diversity of these identities or that race and ethnicity are separate constructs (Helms, 2007). Additionally, calculating ADI using zip code provides a less precise measure of SES than using census block group (Krieger et al., 2003). Despite precedent for using zip code-based ADI to quantify health inequities (e.g., Badreddine et al., 2024; Lehr et al., 2023), future research can examine ED outcomes across other and more individualized measures of disadvantage (Diemer et al., 2013). Finally, as a self-report questionnaire, the EDE-Q is susceptible to inaccuracies in reporting and does not account for all ED presentations, including symptom experiences of marginalized groups (Alexander et al., 2024; Parker et al., 2023).

Historical ED stereotypes and systemic barriers to healthcare have long contributed to inequities in ED outcomes. By examining ED severity and treatment response across several intersecting social groups, the current study preliminarily demonstrates the complexity with which related systems of power and oppression differentially impact the lived experiences of individuals with EDs. Additional research is needed to further describe the illness and treatment trajectories of underrepresented patients, both through the quantitative methods used in this study and qualitative report. Consideration of the variability of patient experiences and perspectives, via an intersectionality framework, is an important step toward providing more equitable care to all those affected by EDs.

## Data Availability

All data produced in the present study are available upon reasonable request to the authors.

## Ethical Standards

The current study was approved by the Salus Institutional Review Board. All participants and their legal guardians provided informed assent and consent.

---

**Figure 1.**
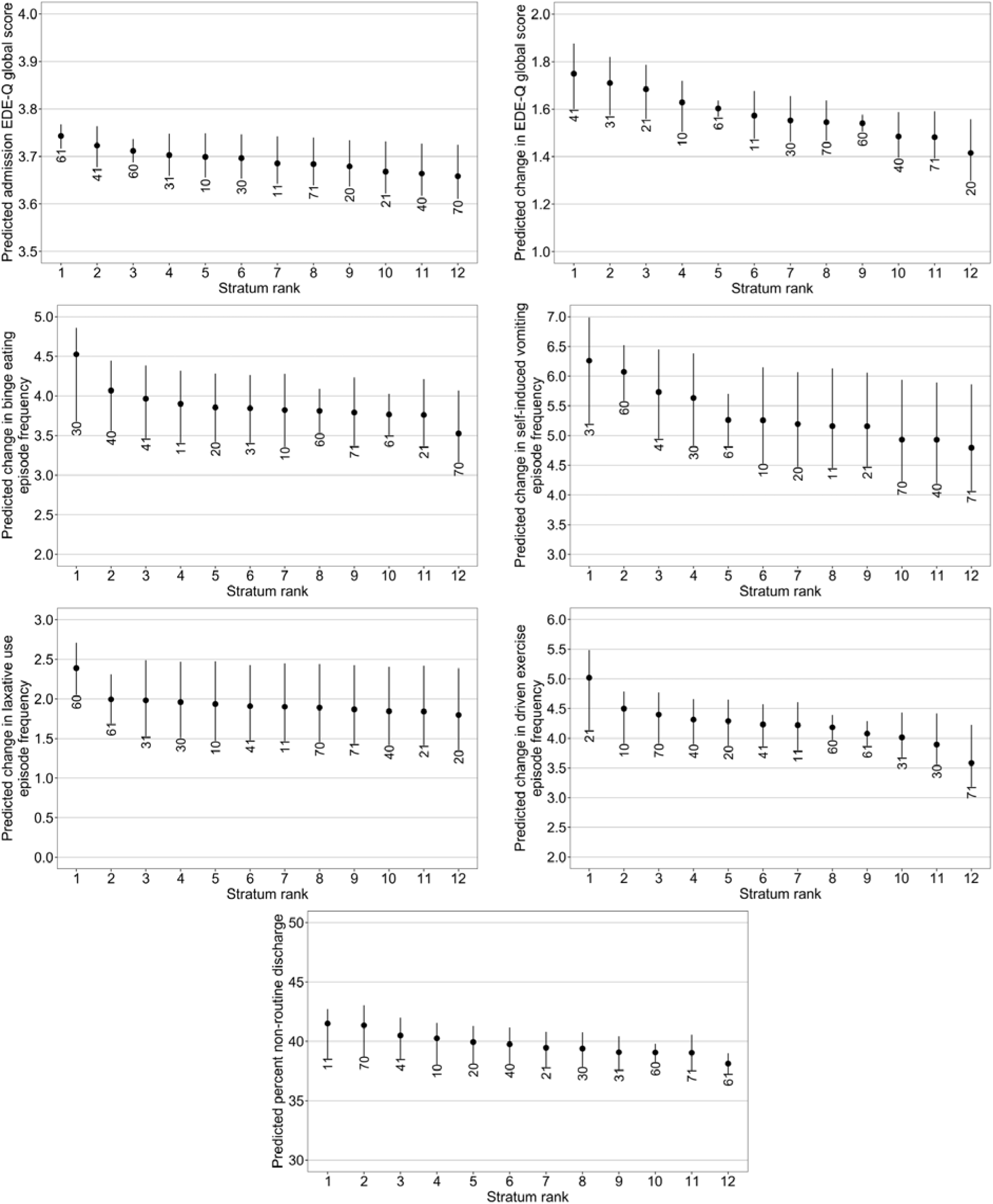
Predicted values for strata defined by race/ethnicity and socioeconomic status. EDE-Q = Eating Disorder Examination-Questionnaire. Spikes indicate approximate 95% confidence intervals. Two-digit stratum ID numbers reflect coding as follows: *Digit 1* = race/ethnicity [1 = Indigenous; 2 = Asian; 3 = Black/African American; 4 = Hispanic/Latine; 6 = White; 7 = Bi/multiracial]. *Digit 2* = socioeconomic status [0 = Higher; 1 = Lower].

**Figure 2.**
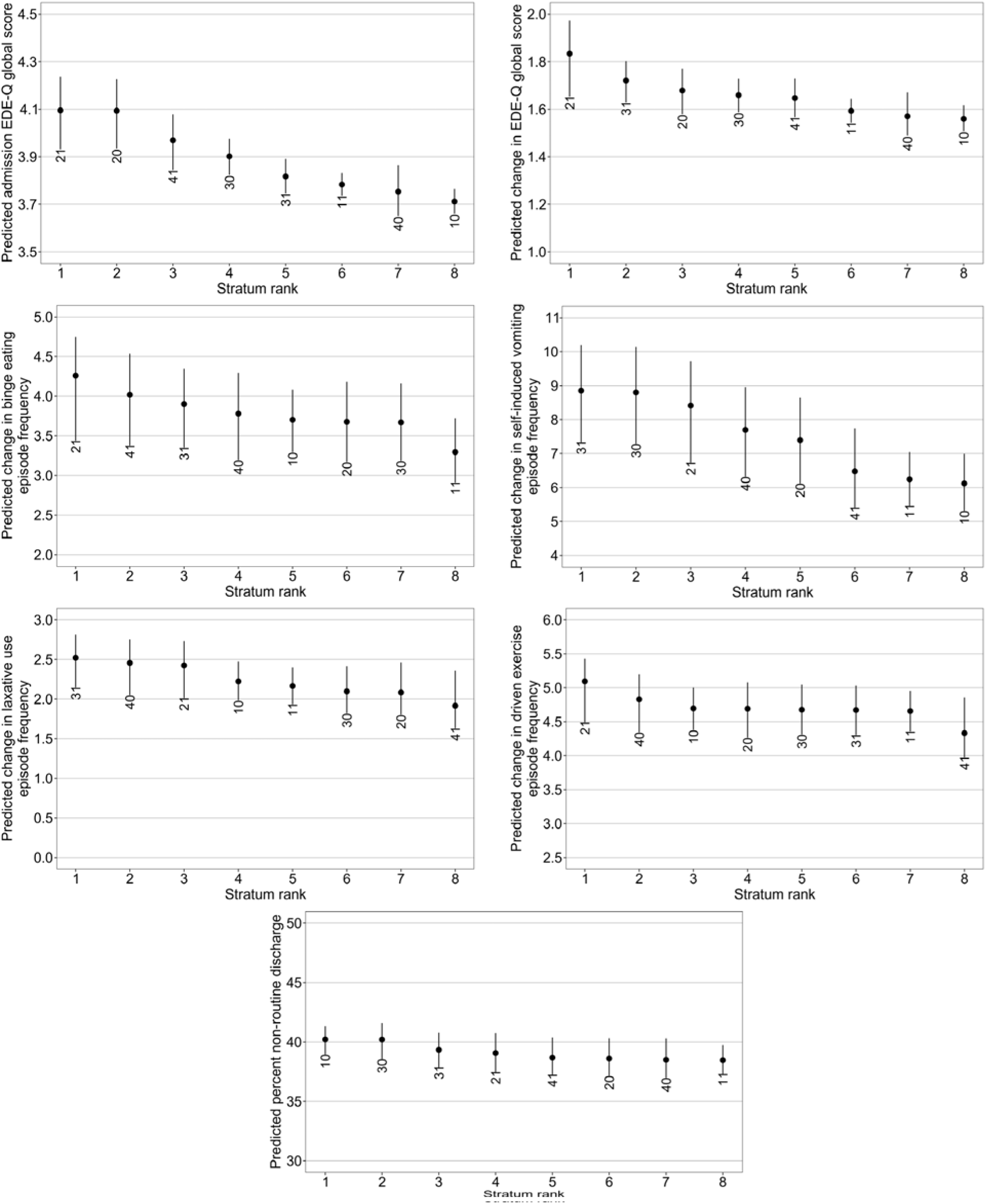
Predicted values for strata defined by sexual orientation and socioeconomic status. EDE-Q = Eating Disorder Examination-Questionnaire. Spikes indicate approximate 95% confidence intervals. Two-digit stratum ID numbers reflect coding as follows: *Digit 1* = sexual orientation [1 = Heterosexual; 2 = Gay or lesbian; 3 = Bisexual, pansexual, or queer; 4 = Other]. *Digit 2* = socioeconomic status [0 = Higher; 1 = Lower].

