## Supplementary Material for "Eating Disorder Severity and Treatment Outcome Across Race/Ethnicity, Sexual Orientation, and Socioeconomic Status: Intersectional Inequities in a Clinical Sample"

**Description of Eating Disorder Treatment**

All treatment sites operated under the same governance structure and utilized a singular eating disorder (ED) treatment approach. The level of care at which patients were admitted (i.e., inpatient [IP], non-hospital-based residential treatment [RES], partial hospitalization program [PHP], or intensive outpatient program [IOP]) was determined based on patients’ degree of medical stability and ED severity. Patients typically stepped down to less-intensive levels of care as their physical and psychological ED symptoms improved, before being discharged to outpatient treatment in the community. In some cases, participants were briefly transferred to more-intensive levels of care (i.e., higher levels of care) before progressing to lower levels of care at the treatment facility. Health insurance coverage also influenced level of care decisions; for example, a patient’s health insurance company might deny coverage after determining that a higher level of care (e.g., IP) was no longer needed. Additionally, some patients’ health insurance plans did not cover RES, thereby requiring a patient to step down from IP to PHP.

At IP, RES, and PHP, patients attended twice-weekly individual psychotherapy sessions, weekly family therapy, twice-weekly psychiatry appointments (daily at IP), medical provider sessions as needed, and weekly sessions with registered dieticians. Medical care was available on site for any medical concerns. Additionally, treatment included three to four hours of evidence-based groups per day, which incorporated interventions based on cognitive behavior therapy, acceptance and commitment therapy, and dialectical behavior therapy. Patients ate three supervised meals and two to three supervised snacks per day. Those in IP and RES resided at the treatment site, while those in PHP resided in the community but attended treatment for 8-10 hours per day, seven days per week. At IOP, treatment consisted of weekly individual psychotherapy, dietary sessions every other week, and 3-4 hours of groups per day, three days per week. Patients in IOP ate one supervised meal each day that they attended treatment. At certain sites, a virtual IOP was also available for patients to attend from home.

**Table S1**

*Sample Sizes Across the Intersections of Race/Ethnicity, Sexual Orientation, and Socioeconomic Status*

| Race/ethnicity | Sexual orientation | SES | *n* |
| --- | --- | --- | --- |
| Indigenous^a^ | Heterosexual | Higher | 2 |
|  |  | Lower | 5 |
|  | Gay or lesbian | Higher | 0 |
|  |  | Lower | 0 |
|  | Bisexual, pansexual, or queer | Higher | 0 |
|  |  | Lower | 2 |
|  | Other^b^ | Higher | 0 |
|  |  | Lower | 0 |
| Asian | Heterosexual | Higher | 12 |
|  |  | Lower | 9 |
|  | Gay or lesbian | Higher | 1 |
|  |  | Lower | 1 |
|  | Bisexual, pansexual, or queer | Higher | 2 |
|  |  | Lower | 5 |
|  | Other | Higher | 3 |
|  |  | Lower | 0 |
| Black/African American | Heterosexual | Higher | 15 |
|  |  | Lower | 15 |
|  | Gay or lesbian | Higher | 1 |
|  |  | Lower | 2 |
|  | Bisexual, pansexual, or queer | Higher | 3 |
|  |  | Lower | 7 |
|  | Other | Higher | 2 |
|  |  | Lower | 3 |
| Hispanic/Latine | Heterosexual | Higher | 19 |
|  |  | Lower | 32 |
|  | Gay or lesbian | Higher | 2 |
|  |  | Lower | 4 |
|  | Bisexual, pansexual, or queer | Higher | 9 |
|  |  | Lower | 8 |
|  | Other | Higher | 3 |
|  |  | Lower | 4 |
| White | Heterosexual | Higher | 387 |
|  |  | Lower | 365 |
|  | Gay or lesbian | Higher | 36 |
|  |  | Lower | 33 |
|  | Bisexual, pansexual, or queer | Higher | 124 |
|  |  | Lower | 105 |
|  | Other | Higher | 31 |
|  |  | Lower | 41 |
| Bi/multiracial | Heterosexual | Higher | 18 |
|  |  | Lower | 16 |
|  | Gay or lesbian | Higher | 3 |
|  |  | Lower | 0 |
|  | Bisexual, pansexual, or queer | Higher | 8 |
|  |  | Lower | 4 |
|  | Other | Higher | 1 |
|  |  | Lower | 0 |

*Note. N* = 3,016. SES = socioeconomic status as measured by area deprivation index (ADI) divided along the median (ADI=82.75).

^a^ Indigenous includes American Indian/Native American or Alaska Native and Native Hawaiian/Pacific Islander participants.

^b^ Other sexual orientation includes participants who identified as asexual, mostly straight, multiple identity, other, and questioning.

**Table S2**

*Parameter Estimates for Baseline-Adjusted Simple Intersectional Models*

|  | Race/ethnicity and SES | | Sexual orientation and SES | |
| --- | --- | --- | --- | --- |
|  | Estimate | [95% CI] | Estimate | [95% CI] |
| Change in EDE-Q global score |  |  |  |  |
| Intercept | -0.17 | [-0.36, 0.06] | -0.16 | [-0.40, 0.10] |
| Baseline EDE-Q global score | 0.47 | [0.43, 0.50] | 0.46 | [0.40, 0.51] |
| Stratum-level variance | 0.14 | [0.01, 0.40] | 0.10 | [0.00, 0.34]] |
| Individual-level variance | 1.19 | [1.15, 1.23] | 1.19 | [1.13, 1.25] |
| VPC | 1.4% |  | 0.7% |  |
| Change in binge eating episode^a^ |  |  |  |  |
| Intercept | -0.73 | [-1.13, -0.31] | -0.58 | [-1.37, 0.31] |
| Baseline binge eating | 0.92 | [0.90, 0.94] | 0.87 | [0.83, 0.91] |
| Stratum-level variance | 0.26 | [0.01, 0.83] | 0.62 | [0.04, 1.64] |
| Individual-level variance | 4.67 | [4.50, 4.84] | 6.16 | [5.86, 6.50] |
| VPC | 0.3% |  | 1.0% |  |
| Change in self-induced vomiting^a^ |  |  |  |  |
| Intercept | -0.37 | [-0.66, -0.08] | -0.28 | [-0.73, 0.18] |
| Baseline self-induced vomiting | 0.94 | [0.94, 0.95] | 0.93 | [0.92, 0.94] |
| Stratum-level variance | 0.19 | [0.01, 0.58] | 0.29 | [0.01, 0.95] |
| Individual-level variance | 3.11 | [3.01, 3.22] | 3.79 | [3.59, 4.00] |
| VPC | 0.4% |  | 0.6% |  |
| Change in laxative use^a^ |  |  |  |  |
| Intercept | -0.23 | [-0.38, -0.06] | -0.17 | [-0.37, 0.00] |
| Baseline laxative use | 0.99 | [0.98, 0.99] | 0.96 | [0.94, 0.98 |
| Stratum-level variance | 0.12 | [0.01, 0.38] | 0.13 | [0.00, 0.41] |
| Individual-level variance | 1.68 | [1.62, 1.74] | 1.69 | [1.61, 1.79] |
| VPC | 0.5% |  | 0.5% |  |
| Change in driven exercise^a^ |  |  |  |  |
| Intercept | -0.43 | [-0.89, 0.20] | -0.54 | [-1.40, 0.20] |
| Baseline driven exercise | 0.78 | [0.75, 0.80] | 0.79 | [0.75, 0.83] |
| Stratum-level variance | 0.34 | [0.01, 1.09] | 0.60 | [0.03, 1.87] |
| Individual-level variance | 4.94 | [4.77, 5.12] | 5.11 | [4.85, 5.38] |
| VPC | 0.5% |  | 1.4% |  |

*Note.* SES = socioeconomic status; EDE-Q = Eating Disorder Examination-Questionnaire; VPC = variance partition coefficient.

^a^ Change in frequency over the past 28 days from admission to discharge.

^b^ 0 = routine discharge and 1 = non-routine discharge.

**Table S3**

*Ranked Strata for Predicted Mean EDE-Q Global Score at Admission*

| Race/ethnicity and SES model | |  |  |  |  |  |
| --- | --- | --- | --- | --- | --- | --- |
| Rank | Race/ethnicity | SES | *n* | Observed *M* | Predicted *M* | Approximate 95% CI |
| 1 | White | Lower | 1262 | 3.77 | 3.74 | [3.72, 3.77] |
| 2 | Hispanic/Latine | Lower | 89 | 3.83 | 3.72 | [3.68, 3.76] |
| 3 | White | Higher | 1247 | 3.71 | 3.71 | [3.69, 3.74] |
| 4 | Black/African American | Lower | 58 | 3.74 | 3.70 | [3.66, 3.75] |
| 5 | Indigenous | Higher | 3 | 4.17 | 3.70 | [3.66, 3.75] |
| 6 | Black/African American | Higher | 37 | 3.70 | 3.70 | [3.65, 3.75] |
| 7 | Indigenous | Lower | 10 | 3.38 | 3.69 | [3.64, 3.74] |
| 8 | Bi/multiracial | Lower | 40 | 3.58 | 3.68 | [3.64, 3.74] |
| 9 | Asian | Higher | 55 | 3.57 | 3.68 | [3.64, 3.73] |
| 10 | Asian | Lower | 39 | 3.43 | 3.67 | [3.62, 3.73] |
| 11 | Hispanic/Latine | Higher | 74 | 3.51 | 3.66 | [3.62, 3.73] |
| 12 | Bi/multiracial | Higher | 64 | 3.45 | 3.66 | [3.61, 3.72] |
| Sexual orientation and SES model | |  |  |  |  |  |
| Rank | Sexual orientation | SES | *n* | Observed *M* | Predicted *M* | Approximate 95% CI |
| 1 | Gay or lesbian | Lower | 42 | 4.39 | 4.10 | [3.93, 4.24] |
| 2 | Gay or lesbian | Higher | 44 | 4.38 | 4.09 | [3.93, 4.23] |
| 3 | Other | Lower | 48 | 4.09 | 3.97 | [3.84, 4.08] |
| 4 | Bisexual, pansexual, or queer | Higher | 149 | 3.92 | 3.90 | [3.82, 3.98] |
| 5 | Bisexual, pansexual, or queer | Lower | 133 | 3.79 | 3.82 | [3.75, 3.89] |
| 6 | Heterosexual | Lower | 452 | 3.77 | 3.78 | [3.74, 3.83] |
| 7 | Other | Higher | 40 | 3.58 | 3.75 | [3.65, 3.87] |
| 8 | Heterosexual | Higher | 462 | 3.68 | 3.71 | [3.66, 3.76] |

*Note*. EDE-Q = Eating Disorder Examination-Questionnaire; SES = socioeconomic status.

**Table S4**

*Ranked Strata for Predicted Change in EDE-Q Global Score from Admission to Discharge*

| Race/ethnicity and SES model | |  |  |  |  |  |  |
| --- | --- | --- | --- | --- | --- | --- | --- |
| Rank | Race/ethnicity | SES | *n* | Observed *M* | Predicted *M* | Approximate 95% CI | Baseline-adjusted predicted *M*  [approximate 95% CI] |
| 1 | Hispanic/Latine | Lower | 38 | 2.05 | 1.75 | [1.60, 1.88] | 1.75 [1.60, 1.85] |
| 2 | Black/African American | Lower | 26 | 2.01 | 1.71 | [1.57, 1.82] | 1.66 [1.57, 1.74] |
| 3 | Asian | Lower | 20 | 1.99 | 1.68 | [1.56, 1.79] | 1.68 [1.57, 1.77] |
| 4 | Indigenous | Higher | 2 | 2.50 | 1.63 | [1.50, 1.72] | 1.63 [1.54, 1.70] |
| 5 | White | Lower | 714 | 1.61 | 1.60 | [1.57, 1.64] | 1.58 [1.55, 1.61] |
| 6 | Indigenous | Lower | 2 | 1.48 | 1.57 | [1.48, 1.68] | 1.64 [1.54, 1.71] |
| 7 | Black/African American | Higher | 11 | 1.44 | 1.55 | [1.46, 1.66] | 1.59 [1.52, 1.66] |
| 8 | Bi/multiracial | Higher | 25 | 1.46 | 1.54 | [1.47, 1.64] | 1.58 [1.51, 1.64] |
| 9 | White | Higher | 671 | 1.53 | 1.54 | [1.51, 1.58] | 1.55 [1.52, 1.58] |
| 10 | Hispanic/Latine | Higher | 42 | 1.33 | 1.48 | [1.40, 1.59] | 1.54 [1.48, 1.61] |
| 11 | Bi/multiracial | Lower | 28 | 1.28 | 1.48 | [1.39, 1.59] | 1.57 [1.51, 1.63] |
| 12 | Asian | Higher | 30 | 1.08 | 1.42 | [1.30, 1.56] | 1.49 [1.41, 1.59] |
| Sexual orientation and SES model | | |  |  |  |  |  |
| Rank | Sexual orientation | SES | *n* | Observed *M* | Predicted *M* | Approximate 95% CI | Baseline-adjusted predicted *M*  [approximate 95% CI] |
| 1 | Gay or lesbian | Lower | 21 | 2.44 | 1.83 | [1.65, 1.97] | 1.69 [1.60, 1.75] |
| 2 | Bisexual, pansexual, or queer | Lower | 83 | 1.82 | 1.72 | [1.63, 1.80] | 1.68 [1.61, 1.73] |
| 3 | Gay or lesbian | Higher | 23 | 1.75 | 1.68 | [1.58, 1.77] | 1.61 [1.56, 1.67] |
| 4 | Bisexual, pansexual, or queer | Higher | 76 | 1.68 | 1.66 | [1.59, 1.73] | 1.61 [1.56, 1.67] |
| 5 | Other | Lower | 32 | 1.65 | 1.65 | [1.57, 1.73] | 1.60 [1.55, 1.66] |
| 6 | Heterosexual | Lower | 261 | 1.57 | 1.59 | [1.54, 1.64] | 1.60 [1.57, 1.64] |
| 7 | Other | Higher | 22 | 1.33 | 1.57 | [1.49, 1.67] | 1.60 [1.56, 1.64] |
| 8 | Heterosexual | Higher | 257 | 1.51 | 1.56 | [1.51, 1.62] | 1.60 [1.56, 1.64] |

*Note*. EDE-Q = Eating Disorder Examination-Questionnaire; SES = socioeconomic status.

**Table S5**

*Ranked Strata for Predicted Change in Binge Eating Episode Frequency from Admission to Discharge*

| Race/ethnicity and SES model | |  |  |  |  |  |  |
| --- | --- | --- | --- | --- | --- | --- | --- |
| Rank | Race/ethnicity | SES | *n* | Observed *M* | Predicted *M* | Approximate 95% CI | Baseline-adjusted predicted *M*  [approximate 95% CI] |
| 1 | Black/African American | Higher | 11 | 15.73 | 4.53 | [3.68, 4.86] | 3.95 [3.77, 4.10] |
| 2 | Hispanic/Latine | Higher | 40 | 5.35 | 4.07 | [3.56, 4.45] | 3.85 [3.70, 4.02] |
| 3 | Hispanic/Latine | Lower | 34 | 4.76 | 3.97 | [3.47, 4.39] | 4.00 [3.80, 4.15] |
| 4 | Indigenous | Lower | 2 | 5.50 | 3.90 | [3.41, 4.32] | 3.91 [3.73, 4.07] |
| 5 | Asian | Higher | 28 | 3.86 | 3.85 | [3.42, 4.28] | 3.91 [3.75, 4.08] |
| 6 | Black/African American | Lower | 25 | 3.92 | 3.84 | [3.41, 4.26] | 3.92 [3.75, 4.08] |
| 7 | Indigenous | Higher | 1 | 0.00 | 3.82 | [3.36, 4.28] | 3.90 [3.72, 4.07] |
| 8 | White | Higher | 622 | 3.80 | 3.81 | [3.54, 4.09] | 3.98 [3.87, 4.08] |
| 9 | Bi/multiracial | Lower | 27 | 3.41 | 3.79 | [3.37, 4.23] | 3.90 [3.74, 4.06] |
| 10 | White | Lower | 675 | 3.70 | 3.76 | [3.51, 4.03] | 3.80 [3.70, 3.91] |
| 11 | Asian | Lower | 18 | 2.83 | 3.76 | [3.36, 4.22] | 3.80 [3.65, 4.00] |
| 12 | Bi/multiracial | Higher | 21 | 0.52 | 3.53 | [3.16, 4.07] | 3.92 [3.75, 4.09] |
| Sexual orientation and SES model | |  |  |  |  |  |  |
| Rank | Sexual orientation | SES | *n* | Observed *M* | Predicted *M* | Approximate 95% CI | Baseline-adjusted predicted *M*  [approximate 95% CI] |
| 1 | Gay or lesbian | Lower | 15 | 10.00 | 4.26 | [3.44, 4.75] | 3.97 [3.51, 4.35] |
| 2 | Other | Lower | 30 | 5.83 | 4.02 | [3.38, 4.53] | 4.01 [3.58, 4.39] |
| 3 | Bisexual, pansexual, or queer | Lower | 76 | 4.50 | 3.90 | [3.35, 4.34] | 4.02 [3.64, 4.34] |
| 4 | Other | Higher | 21 | 4.29 | 3.78 | [3.19, 4.30] | 3.94 [3.51, 4.31] |
| 5 | Heterosexual | Higher | 236 | 3.72 | 3.70 | [3.29, 4.08] | 3.82 [3.58, 4.05] |
| 6 | Gay or lesbian | Higher | 20 | 3.25 | 3.68 | [3.17, 4.18] | 3.84 [3.45, 4.21] |
| 7 | Bisexual, pansexual, or queer | Higher | 68 | 3.60 | 3.67 | [3.18, 4.16] | 3.73 [3.43, 4.02] |
| 8 | Heterosexual | Lower | 248 | 2.80 | 3.29 | [2.91, 3.72] | 3.20 [2.94, 3.48] |

*Note.* SES = socioeconomic status.

**Table S6**

*Ranked Strata for Predicted Change in Self-Induced Vomiting Episode Frequency from Admission to Discharge*

| Race/ethnicity and SES model | |  |  |  |  |  |  |
| --- | --- | --- | --- | --- | --- | --- | --- |
| Rank | Race/ethnicity | SES | *n* | Observed *M* | Predicted *M* | Approximate 95% CI | Baseline-adjusted predicted *M*  [approximate 95% CI] |
| 1 | Black/African American | Lower | 25 | 13.00 | 6.26 | [5.20, 6.99] | 5.31 [5.19, 5.42] |
| 2 | White | Higher | 622 | 6.54 | 6.07 | [5.59, 6.52] | 5.36 [5.28, 5.44] |
| 3 | Hispanic/Latine | Lower | 34 | 8.06 | 5.73 | [4.95, 6.45] | 5.33 [5.21, 5.45] |
| 4 | Black/African American | Higher | 11 | 10.91 | 5.63 | [4.83, 6.39] | 5.31 [5.18, 5.43] |
| 5 | White | Lower | 675 | 5.09 | 5.26 | [4.82, 5.71] | 5.21 [5.13, 5.28] |
| 6 | Indigenous | Higher | 1 | 0.00 | 5.26 | [4.53, 6.15] | 5.29 [5.17, 5.41] |
| 7 | Asian | Higher | 28 | 4.04 | 5.19 | [4.47, 6.07] | 5.32 [5.20, 5.44] |
| 8 | Indigenous | Lower | 2 | 0.00 | 5.16 | [4.45, 6.13] | 5.29 [5.18, 5.41] |
| 9 | Asian | Lower | 19 | 3.79 | 5.16 | [4.48, 6.06] | 5.24 [5.13, 5.37] |
| 10 | Bi/multiracial | Higher | 21 | 1.86 | 4.93 | [4.23, 5.94] | 5.28 [5.16, 5.40] |
| 11 | Hispanic/Latine | Higher | 40 | 2.93 | 4.93 | [4.19, 5.89] | 5.27 [5.16, 5.39] |
| 12 | Bi/multiracial | Lower | 27 | 1.07 | 4.79 | [4.07, 5.86] | 5.31 [5.19, 5.42] |
| Sexual orientation and SES model | | |  |  |  |  |  |
| Rank | Sexual orientation | SES | *n* | Observed *M* | Predicted *M* | Approximate 95% CI | Baseline-adjusted predicted *M*  [approximate 95% CI] |
| 1 | Bisexual, pansexual, or queer | Lower | 76 | 11.51 | 8.85 | [7.34, 10.20] | 7.24 [7.05, 7.39] |
| 2 | Bisexual, pansexual, or queer | Higher | 68 | 11.65 | 8.80 | [7.29, 10.15] | 7.30 [7.09, 7.45] |
| 3 | Gay or lesbian | Lower | 15 | 15.87 | 8.41 | [6.73, 9.72] | 7.10 [6.93, 7.30] |
| 4 | Other | Higher | 21 | 9.90 | 7.70 | [6.32, 8.96] | 7.05 [6.89, 7.25] |
| 5 | Gay or lesbian | Higher | 20 | 8.60 | 7.40 | [6.12, 8.64] | 7.20 [7.00, 7.37] |
| 6 | Other | Lower | 30 | 3.87 | 6.48 | [5.40, 7.74] | 7.16 [6.98, 7.33] |
| 7 | Heterosexual | Lower | 249 | 5.63 | 6.24 | [5.46, 7.04] | 7.02 [6.89, 7.16] |
| 8 | Heterosexual | Higher | 236 | 5.41 | 6.12 | [5.31, 6.99] | 7.13 [7.00, 7.26] |

*Note.* SES = socioeconomic status.

**Table S7**

*Ranked Strata for Predicted Change in Laxative Use Episode Frequency from Admission to Discharge*

| Race/ethnicity and SES model | |  |  |  |  |  |  |
| --- | --- | --- | --- | --- | --- | --- | --- |
| Rank | Race/ethnicity | SES | *n* | Observed *M* | Predicted *M* | Approximate 95% CI | Baseline-adjusted predicted *M*  [approximate 95% CI] |
| 1 | White | Higher | 622 | 2.79 | 2.39 | [2.05, 2.71] | 1.78 [1.74, 1.82] |
| 2 | White | Lower | 675 | 1.95 | 2.00 | [1.69, 2.31] | 1.70 [1.66, 1.75] |
| 3 | Black/African American | Lower | 25 | 2.32 | 1.98 | [1.53, 2.49] | 1.74 [1.68, 1.81] |
| 4 | Black/African American | Higher | 11 | 2.55 | 1.96 | [1.51, 2.47] | 1.73 [1.66, 1.81] |
| 5 | Indigenous | Higher | 1 | 0.00 | 1.94 | [1.47, 2.47] | 1.75 [1.68, 1.82] |
| 6 | Hispanic/Latine | Lower | 34 | 1.56 | 1.91 | [1.48, 2.43] | 1.79 [1.71, 1.85] |
| 7 | Indigenous | Lower | 2 | 0.00 | 1.90 | [1.47, 2.45] | 1.75 [1.67, 1.82] |
| 8 | Bi/multiracial | Higher | 21 | 1.14 | 1.89 | [1.45, 2.44] | 1.77 [1.69, 1.84] |
| 9 | Bi/multiracial | Lower | 27 | 1.15 | 1.87 | [1.43, 2.43] | 1.76 [1.69, 1.83] |
| 10 | Hispanic/Latine | Higher | 39 | 0.95 | 1.84 | [1.42, 2.40] | 1.65 [1.58, 1.75] |
| 11 | Asian | Lower | 19 | 0.37 | 1.84 | [1.41, 2.42] | 1.77 [1.69, 1.84] |
| 12 | Asian | Higher | 28 | 0.25 | 1.80 | [1.35, 2.39] | 1.77 [1.69, 1.83] |
| Sexual orientation and SES model | |  |  |  |  |  |  |
| Rank | Sexual orientation | SES | *n* | Observed *M* | Predicted *M* | Approximate 95% CI | Baseline-adjusted predicted *M*  [approximate 95% CI] |
| 1 | Bisexual, pansexual, or queer | Lower | 76 | 3.30 | 2.52 | [2.15, 2.81] | 1.80 [1.72, 1.86] |
| 2 | Other | Higher | 21 | 4.05 | 2.46 | [2.05, 2.75] | 1.68 [1.60, 1.79] |
| 3 | Gay or lesbian | Lower | 15 | 4.13 | 2.43 | [2.01, 2.73] | 1.77 [1.69, 1.86] |
| 4 | Heterosexual | Higher | 236 | 2.24 | 2.22 | [1.98, 2.47] | 1.77 [1.72, 1.83] |
| 5 | Heterosexual | Lower | 249 | 2.10 | 2.16 | [1.93, 2.40] | 1.75 [1.70, 1.81] |
| 6 | Bisexual, pansexual, or queer | Higher | 68 | 1.76 | 2.10 | [1.82, 2.42] | 1.79 [1.71, 1.86] |
| 7 | Gay or lesbian | Higher | 20 | 1.10 | 2.08 | [1.80, 2.46] | 1.77 [1.69, 1.84] |
| 8 | Other | Lower | 30 | 0.37 | 1.92 | [1.63, 2.36] | 1.68 [1.61, 1.79] |

*Note.* SES = socioeconomic status.

**Table S8**

*Ranked Strata for Predicted Change in Driven Exercise Episode Frequency from Admission to Discharge*

| Race/ethnicity and SES model | |  |  |  |  |  |  |
| --- | --- | --- | --- | --- | --- | --- | --- |
| Rank | Race/ethnicity | SES | *n* | Observed *M* | Predicted *M* | Approximate 95% CI | Baseline-adjusted predicted *M*  [approximate 95% CI] |
| 1 | Asian | Lower | 18 | 10.17 | 5.02 | [4.14, 5.48] | 4.41 [4.08, 4.64] |
| 2 | Indigenous | Higher | 1 | 28.00 | 4.50 | [3.91, 4.79] | 4.30 [4.02, 4.48] |
| 3 | Bi/multiracial | Higher | 21 | 5.62 | 4.40 | [3.91, 4.77] | 4.37 [4.07, 4.56] |
| 4 | Hispanic/Latine | Higher | 40 | 4.73 | 4.31 | [3.90, 4.66] | 4.26 [4.03, 4.44] |
| 5 | Asian | Higher | 28 | 4.79 | 4.29 | [3.88, 4.65] | 4.25 [4.02, 4.43] |
| 6 | Hispanic/Latine | Lower | 34 | 4.41 | 4.23 | [3.87, 4.58] | 4.39 [4.09, 4.60] |
| 7 | Indigenous | Lower | 2 | 6.00 | 4.22 | [3.80, 4.61] | 4.27 [4.01, 4.46] |
| 8 | White | Higher | 622 | 4.21 | 4.19 | [3.98, 4.39] | 4.13 [4.02, 4.25] |
| 9 | White | Lower | 675 | 4.02 | 4.08 | [3.87, 4.29] | 4.06 [3.95, 4.18] |
| 10 | Black/African American | Lower | 25 | 3.24 | 4.02 | [3.68, 4.44] | 4.30 [4.05, 4.48] |
| 11 | Black/African American | Higher | 11 | 1.27 | 3.89 | [3.55, 4.42] | 4.25 [4.00, 4.44] |
| 12 | Bi/multiracial | Lower | 27 | 0.89 | 3.58 | [3.18, 4.23] | 4.06 [3.87, 4.29] |
| Sexual orientation and SES model | |  |  |  |  |  |  |
| Rank | Sexual orientation | SES | *n* | Observed *M* | Predicted *M* | Approximate 95% CI | Baseline-adjusted predicted *M*  [approximate 95% CI] |
| 1 | Gay or lesbian | Lower | 15 | 9.87 | 5.10 | [4.49, 5.43] | 5.45 [4.98, 5.78] |
| 2 | Other | Higher | 21 | 6.05 | 4.83 | [4.34, 5.20] | 4.62 [4.27, 5.08] |
| 3 | Heterosexual | Higher | 236 | 4.74 | 4.70 | [4.38, 5.00] | 5.25 [5.05, 5.45] |
| 4 | Gay or lesbian | Higher | 20 | 5.00 | 4.69 | [4.27, 5.08] | 5.24 [4.88, 5.51] |
| 5 | Bisexual, pansexual, or queer | Higher | 68 | 4.68 | 4.68 | [4.31, 5.05] | 4.99 [4.74, 5.25] |
| 6 | Bisexual, pansexual, or queer | Lower | 76 | 4.70 | 4.67 | [4.30, 5.03] | 5.23 [4.95, 5.50] |
| 7 | Heterosexual | Lower | 248 | 4.61 | 4.66 | [4.36, 4.96] | 4.88 [4.70, 5.08] |
| 8 | Other | Lower | 30 | 2.10 | 4.33 | [3.97, 4.86] | 4.63 [4.28, 5.07] |

*Note.* SES = socioeconomic status.

**Table S9**

*Ranked Strata for Predicted Percentage Non-Routine Discharge*

| Race/ethnicity and SES model | |  |  |  |  |  |
| --- | --- | --- | --- | --- | --- | --- |
| Rank | Race/ethnicity | SES | *n* | Observed % | Predicted % | Approximate 95% CI |
| 1 | Indigenous | Lower | 10 | 80.00 | 41.50 | [38.55, 42.73] |
| 2 | Bi/multiracial | Higher | 64 | 50.00 | 41.35 | [38.75, 43.03] |
| 3 | Hispanic/Latine | Lower | 89 | 43.82 | 40.49 | [38.52, 42.00] |
| 4 | Indigenous | Higher | 3 | 66.67 | 40.25 | [38.05, 41.56] |
| 5 | Asian | Higher | 55 | 41.82 | 39.94 | [38.16, 41.28] |
| 6 | Hispanic/Latine | Higher | 74 | 40.54 | 39.75 | [38.11, 41.15] |
| 7 | Asian | Lower | 39 | 38.46 | 39.46 | [37.81, 40.80] |
| 8 | Black/African American | Higher | 37 | 37.84 | 39.39 | [37.85, 40.76] |
| 9 | Black/African American | Lower | 58 | 36.21 | 39.08 | [37.62, 40.44] |
| 10 | White | Higher | 1247 | 38.97 | 39.06 | [38.26, 39.81] |
| 11 | Bi/multiracial | Lower | 40 | 35.00 | 39.03 | [37.58, 40.55] |
| 12 | White | Lower | 1262 | 37.40 | 38.12 | [37.27, 38.99] |
| Sexual orientation and SES model | |  |  |  |  |  |
| Rank | Sexual orientation | SES | *n* | Observed % | Predicted % | Approximate 95% CI |
| 1 | Heterosexual | Higher | 462 | 41.56 | 40.22 | [38.95, 41.34] |
| 2 | Bisexual, pansexual, or queer | Higher | 149 | 42.95 | 40.21 | [38.55, 41.61] |
| 3 | Bisexual, pansexual, or queer | Lower | 133 | 39.85 | 39.33 | [37.87, 40.77] |
| 4 | Gay or lesbian | Lower | 42 | 38.10 | 39.07 | [37.46, 40.74] |
| 5 | Other | Lower | 48 | 35.42 | 38.68 | [37.24, 40.39] |
| 6 | Gay or lesbian | Higher | 44 | 34.09 | 38.60 | [37.13, 40.32] |
| 7 | Other | Higher | 40 | 32.50 | 38.49 | [36.97, 40.31] |
| 8 | Heterosexual | Lower | 452 | 37.39 | 38.45 | [37.30, 39.73] |

*Note.* SES = socioeconomic status.
